# Common comorbidities in polymyalgia rheumatica and giant cell arteritis: cross-sectional study in UK Biobank

**DOI:** 10.1101/2023.05.08.23289633

**Authors:** Charikleia Chatzigeorgiou, John C. Taylor, Faye Elliott, Eoin P. O’Sullivan, UK Biobank Eye and Vision Consortium, Ann W. Morgan, Jennifer H. Barrett, Sarah L. Mackie

## Abstract

**Objective:** To determine prevalent comorbidities in cases with polymyalgia rheumatica (PMR) or giant cell arteritis (GCA) compared to matched controls.

**Methods:** Nested, cross sectional case-control study within UK Biobank. Case status was defined as self-reported prior diagnosis of PMR or GCA. 10 controls per case were matched for age, sex, ethnicity and assessment centre. Associations with selected self-reported comorbidities were studied using conditional logistic regression.

**Results:** Of PMR (n=1036) or GCA (n=102) cases, 72% were female, 98% white and 58% reported current use of glucocorticoids. Mean age was 63. At the time of the assessment visit, compared to controls, PMR/GCA cases were more likely to report poor general health and at least several days of low mood in the two past weeks. PMR was associated with hypothyroidism (odds ratio (OR) 1.34, 95% confidence interval (CI) 1.07-1.67) and ever-use of hormone replacement therapy (OR 1.26, CI 1.07-1.47). Regarding common comorbidities, PMR and GCA were both associated with hypertension (PMR: OR 1.21, CI 1.06-1.39; GCA: OR 1.86, CI 1.23-2.81) and cataract (PMR: OR 1.51, CI 1.19-1.93; GCA: OR 3.84, CI 2.23-6.60). Additionally GCA was associated with depression (OR 3.05, CI 1.59-5.85). Neither were associated with diabetes.

**Conclusion:** Participants with a history of PMR/GCA, including those not currently taking glucocorticoids, rated their health as poorer than matched controls,. Some previously-described disease associations (hypothyroidism and early menopause) were replicated. Hypertension and cataract, which can both be exacerbated by long-term glucocorticoid therapy, were over-represented in both diseases, particularly GCA.

**Key messages:**

1. Comorbidity was common in individuals with self-reported prior diagnosis of PMR/GCA.
2. PMR/GCA were associated with poorer self-reported health than controls; GCA was associated with depression.
3. Hypertension and cataract were over-represented in PMR/GCA compared with controls but diabetes was not.

## Introduction

Polymyalgia rheumatica (PMR) and giant cell arteritis (GCA) are two overlapping, age-associated inflammatory diseases (1). Both diseases have a peak onset in the mid-70s and a female predominance (2, 3). PMR and GCA are treated with long-term oral glucocorticoid therapy (2, 4). The adverse effects of glucocorticoids include diabetes (5), fracture (4), cardiovascular disease (6) and infection (7). The comorbidity profile of patients with PMR/GCA may be determined by not only the effects of the disease and the effects of glucocorticoid treatment, but also by risk (or protective) factors for PMR/GCA.

Hypertension appears to be a risk factor for both PMR (8) and GCA(9). Conversely, type 2 diabetes has been reported to be protective against both PMR(8) and GCA (10). Furthermore, the literature on cardiovascular risk after a diagnosis of PMR/GCA appears somewhat contradictory (11–13). Studies from the UK Clinical Practice Research Datalink (CPRD) have revealed glucocorticoid dose-response relationships for both cardiovascular disease and infection in PMR/GCA(6, 7), but in another CPRD study, overall mortality was not increased in PMR compared to controls (13). Along similar lines, in a small inception cohort of 359 PMR cases and matched controls, only cataract occurred at greater rates in cases than controls (14). This evidence has been taken as reassurance that the low-to-medium glucocorticoid doses used for PMR are relatively benign compared to the high doses used for GCA.

As regards other risk factors, early menopause has been identified as a risk factor for GCA (15), and a history of multiple pregnancies may possibly be protective (16); it is unclear whether PMR has similar associations.

Current guidelines recommend that GCA should be managed under specialist supervision, whereas PMR is still largely diagnosed and treated in primary care. Mitigating the long-term health and economic impact of GCA and PMR on people and their communities will require a better understanding of the overall needs of patients in both groups.

We sought to explore the multimorbidity burden in people with PMR or GCA, compared to matched controls, in a cross-sectional dataset from the UK Biobank, with a focus on potential GCA/PMR mimics, predisposing factors and glucorticoid-related adverse effects.

## Methods

### Study design and participants

The UK Biobank study is well-described elsewhere (17, 18): 502,649 UK individuals between the ages of 40 and 69 were recruited to a cohort study (further details can be found in the Supplementary Information). At the baseline assessment visit, all participants completed touchscreen questionnaires including sociodemographic factors (e.g. age, sex, ethnicity, postcode of residence), lifestyle (e.g. smoking, alcohol) and self-reported medical history. At the time of the data released under this approval, 20,000 of the original 502,649 participants had had a 5-year repeat assessment. Ethical approval for the UK Biobank study was granted by the North West Multi-Centre Research Ethics Committee, the Community Health Index Advisory Group and the Patient Information Advisory Group (Ref 11/NW/0382) and our study was approved by the UK Biobank ethical review committee (application 5237).

A nested case-control study was conducted. Cases were defined as individuals reporting a prior diagnosis of PMR or GCA at an assessment visit, regardless of whether they reported current oral glucocorticoid therapy at that visit. For those cases who reported having been diagnosed with PMR or GCA at the 5-year assessment visit, but not at the baseline assessment, only data reported at the 5-year assessment were used in this analysis. For controls, data reported during the baseline assessment were used. Each case was matched by age, sex, assessment centre and ethnicity with ten controls who did not report PMR or GCA (further details can be found in the Supplementary information).

We tested for associations of GCA or PMR with overall general health (excellent, good, fair, poor), number of non-cancer illnesses, and self-reported frequency of low mood over the previous two weeks (not at all, several days, more than half the days, or nearly every day). Based on a literature review of potential GCA/PMR predisposing factors/disease associations as well as known glucocorticoid-related adverse effects, we also tested for individual associations with smoking (never/ever), hypothyroidism, average alcohol intake during a typical week over the previous year, and body mass index (BMI). Amongst women, we analysed use of the oral contraceptive pill (OCP, ever/never), number of live births (0, 1, 2, ≥3), history of bilateral oophorectomy, early menopause (<43 years) (15) and use of hormone replacement treatment (HRT) (ever/never). We also examined associations with the three most prevalent self-reported rheumatic diseases (osteoarthritis (OA), rheumatoid arthritis (RA), spondylosis) in PMR/GCA patients.

We tested for associations with self-reported comorbidities particularly relevant to glucocorticoid therapy including hypertension, cataract, diabetes, angina, depression, myocardial infarction, stroke, glaucoma, diabetic eye disease and transient ischaemic attack. In the subset of cases with available data on ocular measurements (Appendix 1) we also compared intraocular pressure (IOP) and visual acuity.

There are potential inaccuracies in self-reported PMR or GCA status. PMR especially is well-recognised to be mimicked by several other conditions potentially leading to diagnostic confusion (19, 20). However, since comorbidity is common in the age group affected by PMR (21), the coexistence of a comorbidity with PMR does not necessarily prevent or invalidate the diagnosis of PMR. We created a list of 21 candidate mimicking conditions (Supplementary Table 3), selected *a priori* from clinical experience, our prior work (22) and a literature review, and explored their frequency in cases compared to controls. This was used to inform a sensitivity analysis of “isolated PMR”, in which we excluded the PMR cases with co-existent mimicking conditions that occurred with >1% frequency in the PMR group and showed a significant association with PMR.

### Statistical analysis

Associations were assessed with frequency tables and Pearson’s chi-squared tests for two independent proportions. Mean and 95% confidence interval (CI) were calculated to summarize continuous variables and were compared by t-tests. Conditional logistic regression was used to study the association between case-control status and potentially related variables. For categorical variables, the most prevalent category among both cases and controls was chosen as the reference category. Candidate comorbidities with a frequency of at least 1.0% were analyzed further using conditional logistic regression to test for case-control differences (significance level 5%).

For the subset that had detailed ocular measurements at their assessment centre visit, to take account of separate measurements being made for each eye, random-effect models were used to compare intraocular pressure (IOP) and visual acuity in PMR cases reporting current glucocorticoid use, compared to PMR cases not reporting current glucocorticoid use at the time of the assessment.

As the proportion of missing data in the explored variables was 5% or less, no imputation was carried out for missing data (23). Statistical analysis was performed with Stata 14.0 statistical software (StataCorp. 2015. *Stata Statistical Software: Release 14*. College Station, TX: StataCorp LP).

## Results

1036 cases with PMR and 102 cases with GCA, including 23 cases with both conditions, were identified from the self-reported diagnostic codes (Supplementary Figure 1). 72% were female and 98% were classified as White. Assuming a 5% significance level and an exposure prevalence of 20%, these sample sizes give 99% and 31% power, respectively, to detect an odds ratio of 1.4 in a 1:10 matched sample (Supplementary Tables 1-2).

At the time of the assessment visit, mean age of PMR/GCA cases was 63 years (median 64 years; interquartile range 61, 67 years), and 58% reported currently taking oral glucocorticoid therapy. Forty-one PMR cases (4.0%) and 5 GCA cases (4.9%) reported taking methotrexate. Tocilizumab was not approved for GCA at the time of the assessment visits in our dataset.

Comorbidity was common. The proportion of patients reporting two or more, and four or more non-cancer illnesses was, respectively, 66% and 31% of PMR cases (Table 4) and 86% and 46% of those with GCA (Supplementary Table 5).

### Potential PMR/GCA mimics

First, we explored the data for co-diagnosis of other conditions that could mimic PMR or GCA. Three of these conditions, OA, RA and spine arthritis/cervical spondylosis, were found to occur with a frequency > 1% (Supplementary Table 3). Of the PMR cases, 38 reported a diagnosis of RA and 168 reported a diagnosis of OA. We found possible associations of each of these potential mimics with both PMR and GCA; as expected, the evidence for an association between RA and PMR was strongest (OR:2.08, CI:1.46-2.98), p=5.1 × 10^−5^), although there was also a statistically significant association between OA and PMR (OR:1.37, CI:1.14-1.63), p=0.001); there was no clear association between spine arthritis and PMR (OR:1.51, CI:1.00-2.27), p=0.05) (Table 1). Many of the RA diagnoses came shortly after the PMR diagnosis, although there was a minority of patients who reported being diagnosed with RA substantially before the PMR diagnosis (Supplementary Figures 2, 3). Therefore, in subsequent analyses we also conducted sensitivity analyses restricted to “isolated PMR” (without GCA, OA or RA). The number of GCA cases was too small to perform these sensitivity analyses.

**Table 1.** Associations of PMR and GCA with potential PMR or GCA mimics.

|  | **PMR dataset**<br>PMR cases (n=1036) vs CTRL (n=10360) |  |  |  | **GCA dataset**<br>GCA cases (n=102) vs CTRL (n=1020) |  |  |  |
| --- | --- | --- | --- | --- | --- | --- | --- | --- |
|  | PMR | CTRL | OR (CI) | P | GCA | CTRL | OR (CI) | P |
| Osteoarthritis | 168<br>(16.2%) | 1293<br>(12.5%) | 1.37<br>(1.14-1.63) | 0.001 | 19<br>(18.6%) | 120<br>(11.8%) | 1.73<br>(1.01-2.98) | 0.046 |
| Rheumatoid arthritis | 38<br>(3.7%) | 186<br>(1.8%) | 2.08<br>(1.46-2.98) | 5.1 x<br>10<sup>-05</sup> | <5 | 12<br>(1.2%) | 3.42<br>(1.08-10.8) | 0.036 |
| Spine arthritis/<br>Cervical<br>spondylosis | 27<br>(2.6%) | 181<br>(1.75%) | 1.51<br>(1.00-2.27) | 0.05 | 5<br>(4.9%) | 14<br>(1.4%) | 3.6<br>(1.30-10.3) | 0.01 |

OR: odds ratio from conditional regression analysis; SD: Standard deviation; CI: 95% Confidence interval; P: p-value from the conditional logistic regression; For this analysis cases were matched with controls by age, sex, ethnicity and assessment centre; n: sample size; CTRL: controls, matched for age, sex, ethnicity and assessment centre; For this analysis cases were matched with controls by age, sex, ethnicity and assessment centre

### Candidate predisposing factors

Examining candidate predisposing factors (Table 2), we did not find any association of smoking with PMR or GCA. There was no clear association with frequency of alcohol consumption for PMR, but GCA cases were more likely to report never drinking in the previous year compared to the most common category of once or twice per week (OR:3.55, CI:1.82-6.89), p=1.9×10^−4^). We found a significant association between hypothyroidism and PMR (OR:1.34, CI:1.07-1.67, p=0.009), which was not markedly attenuated in the subset with “isolated PMR”. The number of cases with GCA was too small to confirm or exclude an association with hypothyroidism.

**Table 2.**
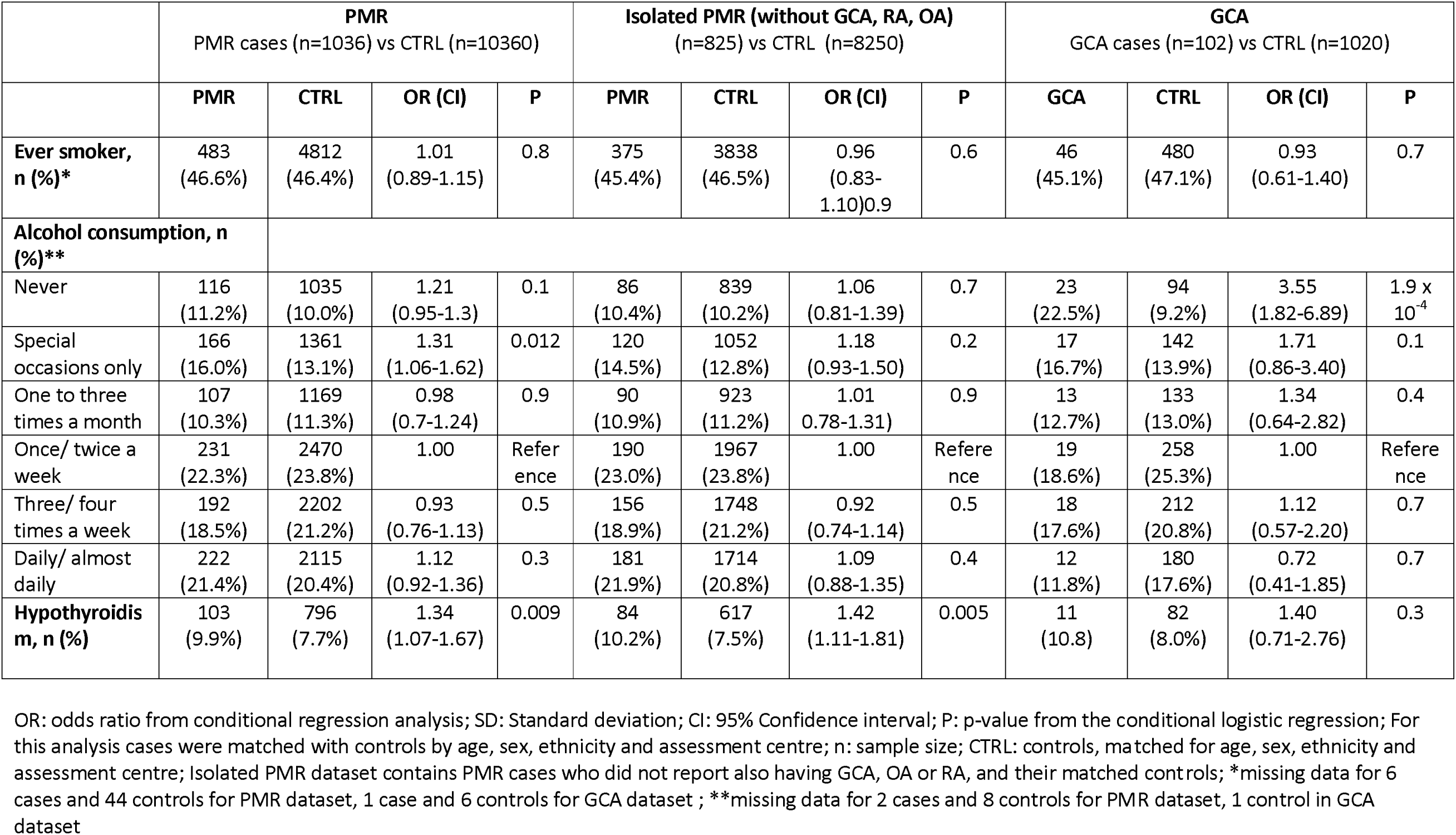
Candidate PMR and GCA susceptibility factors.

Within females (Table 3), PMR was associated with ever-use of HRT, compared to never-use (OR:1.26, CI:1.07-1.47, p=0.005), with a similar effect size observed in the “isolated PMR” sensitivity analysis. A possible association of GCA was seen with early menopause (OR:1.90, CI:1.01-3.59, p=0.047). Neither PMR nor GCA were associated with the other susceptibility factors considered, including number of live births, prior use of oral contraceptive pill or history of bilateral oophorectomy.

**Table 3.** Candidate PMR and GCA susceptibility factors specific to females.

|  | **PMR**<br>PMR cases (n=747) vs CTRL (n=7470) |  |  |  | **Isolated PMR (without GCA, RA, OA)**<br>(n=574) vs CTRL (n=5740) |  |  |  | **GCA**<br>GCA cases (n=73) vs CTRL (n=730) |  |  |  |
| --- | --- | --- | --- | --- | --- | --- | --- | --- | --- | --- | --- | --- |
|  | PMR | CTRL | OR (CI) | P | PMR | CTRL | OR (CI) | P | GCA | CTRL | OR (CI) | P |
| **Number of live births\*** |  |  |  |  |  |  |  |  |  |  |  |  |
| 0 | 109<br>(14.6%) | 1069<br>(14.3%) | 1.02<br>(0.81-1.29) | 0.8 | 88<br>(15.3%) | 853<br>(14.9%) | 1.06<br>(0.82-1.36) | 0.7 | 14<br>(19.2%) | 94<br>(12.9%) | 1.55<br>(0.79-3.02) | 0.2 |
| 1 | 79<br>(10.6%) | 865<br>(11.6%) | 0.92<br>(0.71-1.19) | 0.5 | 68<br>(11.8%) | 643<br>(11.2%) | 1.08<br>(0.82-1.44) | 0.6 | 6<br>(8.2%) | 87<br>(11.9%) | 0.73<br>(0.29-1.83) | 0.5 |
| 2 | 339<br>(45.4%) | 3412<br>(45.7%) | 1.00 | Reference | 255<br>(44.4%) | 2616<br>(45.6%) | 1.00 | Reference | 33<br>(45.2%) | 347<br>(47.5%) | 1.00 | Reference |
| >=3 | 219<br>(29.3%) | 2119<br>(28.4%) | 1.04<br>(0.87-1.24) | 0.7 | 162<br>(28.2%) | 1625<br>(28.3%) | 1.02<br>(0.83-1.26) | 0.8 | 20<br>(27.4%) | 201<br>(27.5%) | 1.05<br>(0.58-1.90) | 0.9 |
| **Menopause <43 years, n (%)** * | 66 (8.8%) | 536<br>(7.2%) | 1.28<br>(0.97-1.70) | 0.08 | 49<br>(8.5%) | 405<br>(7.1%) | 1.25<br>(0.91-1.73) | 0.2 | 16<br>(21.9%) | 101<br>(13.8%) | 1.90<br>(1.01-3.59) | 0.047 |
| **Ever received menopause hormone therapy, n (%)** | 445<br>(59.6%) | 4063<br>(54.4%) | 1.26<br>(1.07-1.47) | 0.005 | 328<br>(57.1%) | 3061<br>(53.3%) | 1.19<br>(0.99-1.42) | 0.06 | 45<br>(61.6%) | 404<br>(55.3%) | 1.30<br>(0.79-2.15) | 0.3 |
| **History of bilateral oophorectomy, n (%)** | 87<br>(11.6%) | 801<br>(10.7%) | 1.11<br>(0.88-1.41) | 0.4 | 58<br>(10.1%) | 612<br>(10.7%) | 0.95<br>(0.71-1.26) | 0.7 | 11<br>(15.1%) | 75<br>(10.3%) | 1.51<br>(0.76-3.00) | 0.2 |
| **Ever taken oral contraceptive pills, n (%)** | 538<br>(72.0%) | 5349<br>(71.6%) | 1.02<br>(0.86-1.22) | 0.8 | 413<br>(72.0%) | 4125<br>(71.9%) | 1.01<br>(0.83-1.23) | 0.9 | 51<br>(69.9%) | 524<br>(71.8%) | 0.90<br>(0.52-1.55) | 0.7 |

OR: odds ratio from conditional regression analysis; SD: Standard deviation; CI: 95% Confidence interval; P: p-value from the conditional logistic regression; For this analysis cases were matched with controls by age, sex, ethnicity and assessment centre; n: sample size; CTRL: controls, matched for age, sex, ethnicity and assessment centre; For this analysis cases were matched with controls by age, sex, ethnicity and assessment centre; Isolated PMR dataset contains PMR cases who did not report also having GCA, OA or RA, and their matched controls; \*Variables with this label contain missing data <5%

For PMR cases also reporting a diagnosis of hypertension, 67% of cases reported an earlier onset of hypertension than the onset of PMR (Supplementary Figure 3). We considered whether hypothyroidism or hypertension might have been identified during the diagnostic workup for PMR. Although this possibility could not be excluded from the data we had, cases with PMR reported that they were diagnosed with hypothyroidism a mean of around 10 years before they were diagnosed with PMR (Supplementary Figures 2,3).

### Self-rating of general health and recent low mood

Both PMR (Table 4) and GCA (Supplementary Table 5) cases were more likely to rate their general health as poor than matched controls. Both PMR and GCA (Supplementary Tables 4 and 5) cases were more likely to report low mood for at least several days during the previous 2 weeks. Similar results were observed in sensitivity analysis of isolated PMR cases, and also even when considering only isolated PMR cases no longer taking glucocorticoid therapy (Supplementary Table 4).

**Table 4:**
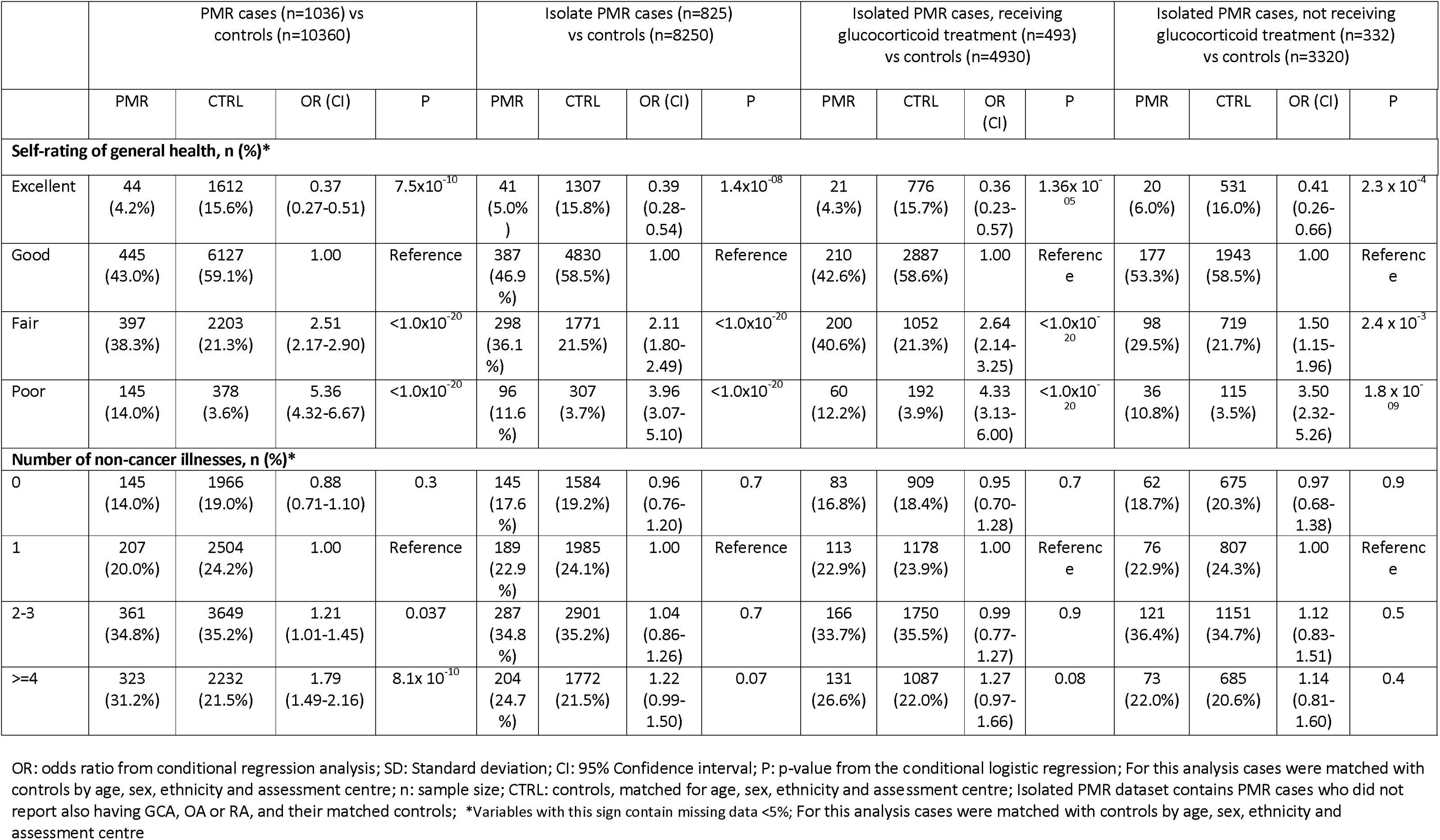
General health of patients with PMR compared to matched controls.

### Comorbidities relevant to glucocorticoid therapy

Both PMR and GCA were significantly associated with hypertension (PMR: OR 1.21, CI 1.06-1.39, Table 5; GCA: OR 1.86, CI 1.23-2.81, Table 6). In the subset of isolated PMR patients no longer receiving glucocorticoid therapy, similar frequencies of hypertension were observed in cases as in controls (Table 5).

**Table 5.** Comorbidities relevant to glucocorticoid therapy in PMR, with sensitivity analyses.

|  | PMR cases vs CTRL |  |  |  | Isolated PMR cases vs CTRL |  |  |  | Isolated PMR, **receiving glucocorticoid** treatment vs CTRL |  |  |  | Isolated PMR, **not receiving glucocorticoid** treatment vs CTRL |  |  |  |
| --- | --- | --- | --- | --- | --- | --- | --- | --- | --- | --- | --- | --- | --- | --- | --- | --- |
|  | PMR<br>(n=1036) | CTRL<br>(n=10360) | OR (CI) | P | PMR<br>(n=825) | CTRL<br>(n=8250) | OR (CI) | P | PMR<br>(n=493) | CTRL<br>(n=4930) | OR (CI) | P | PMR<br>(n=332) | CTRL<br>(n=3320) | OR (CI) | P |
| Hypertension, n (%) | 380<br>(36.7%) | 3354<br>(32.4%) | 1.21<br>(1.06-1.39) | 0.005 | 287<br>(34.8%) | 2683<br>(32.5%) | 1.11<br>(0.95-1.29) | 0.2 | 184<br>(37.3%) | 1614<br>(32.7%) | 1.23<br>(1.01-1.50) | 0.04 | 103<br>(31.0%) | 1069<br>(32.2%) | 0.95<br>(0.74-1.21) | 0.7 |
| Diabetes, n (%) | 50<br>(4.8%) | 526<br>(5.1%) | 0.95<br>(0.70-1.28) | 0.7 | 41<br>(5.0%) | 435<br>(5.3%) | 0.94<br>(0.67-1.31) | 0.7 | 24<br>(4.9%) | 269<br>(5.5%) | 0.88<br>(0.57-1.36) | 0.6 | 17<br>(5.1%) | 166<br>(5.0%) | 1.02<br>(0.61-1.72) | 0.9 |
| Diabetes eye disease, n (%) | 10<br>(1.0%) | 89<br>(0.9%) | 1.12<br>(0.58-2.17) | 0.7 | 5<br>(0.6%) | 82<br>(1.0%) | 0.61<br>(0.25-1.50) | 0.3 | <5 | 46<br>(0.9%) | 0.65<br>(0.20-2.10) | 0.5 | <5 | 36<br>(1.1%) | 0.55<br>(0.13-2.31) | 0.4 |
| Glaucoma, n (%) | 25<br>(2.4%) | 238<br>(2.3%) | 1.05<br>(0.69-1.60) | 0.8 | 21<br>(2.5%) | 182<br>(2.2%) | 1.16<br>(0.73-1.84) | 0.5 | 15<br>(3.0%) | 106<br>(2.1%) | 1.43<br>(0.82-2.50) | 0.2 | 6<br>(1.8%) | 76<br>(2.3%) | 0.78<br>(0.34-1.82) | 0.6 |
| Cataract, n (%) | 85<br>(8.2%) | 583<br>(5.6%) | 1.51<br>(1.19-1.93) | 0.001 | 64<br>(7.8%) | 465<br>(5.6%) | 1.42<br>(1.08-1.87) | 0.01 | 39<br>(7.9%) | 259<br>(5.2%) | 1.56<br>(1.10-2.23) | 0.01 | 25<br>(7.5%) | 206<br>(6.2%) | 1.24<br>(0.80-1.92) | 0.3 |
| Depression, n (%) | 61<br>(5.9%) | 545<br>(5.3%) | 1.13<br>(0.86-1.48) | 0.4 | 43<br>(5.2%) | 444<br>(5.4%) | 0.97<br>(0.70-1.33) | 0.8 | 20<br>(4.1%) | 272<br>(5.5%) | 0.72<br>(0.45-1.15) | 0.2 | 23<br>(6.9%) | 172<br>(5.2%) | 1.36<br>(0.87-2.14) | 0.2 |
| Angina n (%) | 60<br>(5.8%) | 417<br>(4.0%) | 1.42<br>(1.07-1.90) | 0.016 | 41<br>(5.0%) | 330<br>(4.0%) | 1.26<br>(0.90-1.77) | 0.2 | 25<br>(5.1%) | 204<br>(4.1%) | 1.24<br>(0.81-1.92) | 0.3 | 16<br>(4.8%) | 126<br>(3.8%) | 1.29<br>(0.75-2.21) | 0.3 |
| Transient ischaemic attack, n% | 13<br>(1.3%) | 58<br>(0.6%) | 2.46<br>(1.34-4.52) | 0.004 | 6<br>(0.7%) | 37<br>(0.4%) | 1.63<br>(0.68-3.86) | 0.3 | <5 | 20<br>(0.4%) | 2.01<br>(0.68-5.94) | 0.2 | <5 | 17<br>(0.5%) | 1.18<br>(0.27-5.09) | 0.8 |
| Stroke, n (%) | 21<br>(2.0%) | 181<br>(1.7%) | 1.11<br>(0.70-1.77) | 0.6 | 15<br>(1.8%) | 147<br>(1.8%) | 1.02<br>(0.60-1.74) | 0.08 | 9<br>(1.8%) | 85<br>(1.7%) | 1.06<br>(0.53-2.12) | 0.9 | 6<br>(1.8%) | 62<br>(1.9%) | 0.97<br>(0.42-2.24) | 0.9 |

OR: odds ratio from conditional regression analysis; SD: Standard deviation; CI: 95% Confidence interval; P: p-value from the conditional logistic regression; For this analysis cases were matched with controls by age, sex, ethnicity and assessment centre; n: sample size; CTRL: controls, matched for age, sex, ethnicity and assessment centre; For this analysis cases were matched with controls by age, sex, ethnicity and assessment centre

**Table 6:** Comorbidities relevant to glucocorticoid therapy in patients with giant cell arteritis.

|  | **GCA,**<br>**n=102** | **CTRL,**<br>**n=1020** | **OR (CI)** | **P** |
| --- | --- | --- | --- | --- |
| Depression, n (%) | 13 (12.7%) | 46 (4.5%) | 3.05 (1.59-5.85) | 0.001 |
| Hypertension, n (%) | 50 (49.0%) | 349 (34.2%) | 1.86 (1.23-2.81) | 0.003 |
| Diabetes, n (%) | 10 (9.8%) | 60 (5.9%) | 1.75 (0.86-3.57) | 0.1 |
| Diabetic eye disease, n (%) | <5 | 9 (0.88%) | 4.89 (1.41-16.97) | 0.012 |
| Glaucoma, n (%) | 5 (4.9%) | 21 (2.1%) | 2.45 (0.90-6.62) | 0.08 |
| Cataract, n (%) | 20 (19.6%) | 68 (6.7%) | 3.84 (2.23-6.60) | $9.8 \times 10^{-6}$ |
| Angina, n (%) | 7 (6.9%) | 55 (5.4%) | 1.31 (0.57-3.01) | 0.5 |
| Stroke, n (%) | <5 | 20 (2.0%) | 2.03 (0.68-6.04) | 0.2 |
| Transient ischaemic attack, n (%) | <5 | 6 (0.6%) | 7.32 (3.94-27.5) | 0.003 |
| Myocardial infarction, n (%) | <5 | 29 (2.8%) | 0.68 (0.15-2.92) | 0.6 |

OR: odds ratio from conditional regression analysis; SD: Standard deviation; CI: 95% Confidence interval; P: p-value from the conditional logistic regression; CTRL: controls, matched for age, sex, ethnicity and assessment centre; For this analysis, cases were matched with controls with the same age, sex, ethnic background and place of residence

Body mass index (BMI) did not differ significantly between PMR/GCA cases and controls (data not shown). Neither PMR nor GCA was significantly associated with diabetes (Table 5, Table 6); GCA was significantly associated with diabetic eye disease (OR 4.89, CI 1.41-16.97) but the frequency of diabetic eye disease was low giving only very small numbers for the comparison (Table 6).

Both PMR and GCA were significantly associated with cataract (PMR: OR 1.51, CI 1.19-1.93 (Table 5); GCA: OR 3.84, CI 2.23-6.60 (Table 6)). Both PMR and GCA were also significantly associated with transient ischaemic attack, but the frequency of transient ischaemic attack was low giving only very small numbers for the comparison (Tables 5, 6).

GCA was significantly associated with depression (OR: 3.05, CI:1.59-5.85), p=0.001, Table 6).

### Ocular measurements

No significant difference was seen in ocular measurements comparing cases and controls (data not shown). Comparing the 157 PMR cases currently treated with glucocorticoids and the 108 PMR cases not currently treated with glucocorticoids (Supplementary Table 6), and excluding PMR cases with concomitant GCA, glucocorticoid treatment was associated with a significantly higher intraocular pressure (p=0.005; intra class correlation, rho=0.66) but the overall magnitude of the difference was of questionable clinical significance.

## Discussion

In the context of the known multi-system adverse event profile of glucocorticoid therapy, and yet absence of a mortality signal in PMR population-based data and a previously-postulated inverse relationship of GCA with diabetes, we sought to describe the comorbidity associated with a prior diagnosis of PMR and GCA. We found that comorbidity was common in these patients. Two of the commonest comorbidities considered, hypertension and cataract, were over-represented in both diseases compared to controls matched for age, sex, ethnicity and assessment centre. We did not detect a significant association of diabetes with either disease. Participants with PMR/GCA were more likely to rate their general health as poor, and to report low mood for at least several days over the past two weeks. Self-report of GCA was associated with self-report of a depression diagnosis. These results underline the importance of optimising physical and mental health in people with these diagnoses, even after stopping glucocorticoids.

One major limitation of this study was the reliance on self-report of GCA or PMR, and lack of information about historic glucocorticoid treatment; medication information was only available in relation to the time of the assessment visit. At the time this study was done, health record linkage data were not available, and therefore validation of self-report diagnosis was not possible. Particularly for those patients not reporting oral glucocorticoid therapy at the time of the assessment visit, it is possible that GCA/PMR self-report could be inaccurate. Interestingly we found a strong association between PMR and RA, in line with earlier observations by ourselves and others that diagnostic revision can occur between PMR and RA as the clinical picture evolves over time (22) as well as the potential for shared genetic susceptibility (24, 25). Apparent associations might arise from misattribution of the cause of musculoskeletal pain; PMR/GCA can cause arthralgia, but OA, spine arthritis and cervical spondylosis are common causes of musculoskeletal pain in the community. Other possibilities may include body pain resulting from glucocorticoid-related myopathy or other effects of disease or treatment; or flares of OA appearing as part of the glucocorticoid withdrawal syndrome. It is not possible to distinguish between these possibilities with the data we had. However, we used the observed associations to design sensitivity analyses for “isolated PMR” without GCA, RA or OA.

Contrary to previous reports (15, 26) we did not find a clear association with smoking. Our observed data on alcohol intake are difficult to interpret as individuals with various medical conditions may often choose to reduce alcohol intake as part of a self-management approach. However, a previous prospective epidemiological study had reported that individuals with lower prior alcohol intake were more likely to develop PMR subsequently (27). Further research in longitudinal studies is required.

GCA has been previously described as associated with early menopause (15), and we also found a statistically significant association between GCA and early menopause, albeit with small numbers. This may be biologically plausible given the impact of menopause on vascular health. Menopause has multiple health impacts (both systemic inflammation and cardiometabolic effects), that might plausibly influence risk of PMR and GCA. A previous report (28) had suggested that multiple pregnancies are protective, but we did not observe any such association in our dataset.

PMR was associated with prior use of HRT. We were unable to determine the explanation for this: possibilities might include the effects of the systemic inflammatory syndrome of menopause, diagnostic confusion with menopause-related arthralgia/diaphoresis, or use of HRT as part of a strategy to prevent osteoporosis, but the explanations for this might be multifactorial, including early menopause as a possible predisposing factor, or prescription of HRT as part of a therapeutic strategy for prevention of glucocorticoid-induced osteoporosis.

An association between hypothyroidism and PMR has been reported before (8, 29). Guidelines advocate routine thyroid function testing in the diagnostic workup of PMR (30) which could introduce detection bias. The reported dates of the diagnoses of these two conditions was, however, not consistent with this as sole explanation. Shared susceptibility factors for autoimmune diseases (polyautoimmunity) are an alternative explanation: there are well-known associations of thyroid disease with RA (31, 32) and multiple other autoimmune diseases. It has been debated whether PMR should be considered predominantly an autoimmune or an autoinflammatory disease (33); other authors have conceptualized PMR and GCA as two diseases along a common spectrum(1). Eludication of shared and distinct risk factors could help to advance these debates.

We did not find a strong association of PMR/GCA with either smoking or BMI. In line with previous studies(8, 34), we did not find a significant association of diabetes with PMR/GCA, despite the glucocorticoid burden of treatment. In contrast both PMR and GCA were found to be associated with hypertension. This is in line with previous literature (8, 35), but in our cross-sectional dataset interpretation is complicated by difficulty unravelling whether the association with hypertension is due to shared susceptibility factors or disease mechanisms, to detection bias in the workup of PMR/GCA, or to glucocorticoid therapy. Regardless, hypertension was very common in both PMR and GCA; further research is needed to determine optimal hypertension management strategies.

Some comorbidities of potential concern for glucocorticoid therapy were over-represented in PMR/GCA compared to controls, notably cataract. This seems most likely to relate to glucocorticoid therapy. Also notably, depression was associated with GCA and self-reported low mood with both PMR and GCA. Mental health impact is an important part of the comorbidity burden faced by this patient group.

Other than the case ascertainment issues with self-report data, this study also has limitations to generalizability. The UK Biobank cohort is not statistically representative of the UK population (UK Biobank participants have in general higher levels of education and socio-economic status than the general population). Also, the age range of UK Biobank participants at baseline is somewhat younger than the typical age of PMR/GCA onset. These issues could have introduced selection biases and also could have underestimated the comorbidity burden of PMR/GCA: age is a risk factor not only for GCA-related blindness (36–39) but also for most glucocorticoid-related adverse effects (40).

In this study we explored the multimorbidity burden of UK Biobank participants who self-reported diagnoses of PMR or GCA. Comorbidities, including those of concern for glucocorticoid prescribing, were common; there was no association with diabetes or BMI but there was an association with OA. PMR/GCA cases had poorer self-rated health, more frequent low mood, and a higher prevalence of hypertension and cataract than matched controls. Many of these effects were seen even in those who were no longer taking glucocorticoids.

This study contributes to the body of knowledge on the multimorbidity burden in PMR/GCA. An important message for clinicians is to pay attention to indices of vascular health and mental health, and to consider the potential late effects of disease and treatment even when they are no longer taking glucocorticoids.

## Supporting information

Appendix 1 UK GCA Consortium Members

## Acknowledgements

This research was conducted using the UK Biobank Resource under Application Number 5237. We would like to thank UK Biobank participants, without whom this study would not have been possible. The authors thank Mrs. Louise Sorensen for her contribution.

## Funding

Chatzigeorgiou: PhD was supported by a Emma and Leslie Reid Scholarship from the University of Leeds

Barrett: received salary support from the NIHR Leeds BRC

Morgan: has received salary support from the MRC, NIHR Leeds BRC and NIHR Leeds Medtech and *in vitro* Diagnostics Co-operative; NIHR Senior Investigator Award

Mackie: NIHR Clinician Scientist Fellowship NIHR-CS-012-016.

Other authors:

Costs for the data extract for this study were supported by a grant to Eoin O’Sullivan from the Fight for Sight Small Grants Scheme (Grant Award 1477/8). This study was also supported by the National Institute for Health Research (NIHR) Biomedical Research Centre and the NIHR Medtech and In vitro Diagnostics Co-operative. The views expressed are those of the authors and not necessarily those of the NIHR or the Department of Health and Social Care.

## Data availability statement

The data used to support the findings of this study are included within the article.

## Author Contributions

Design, concept: CC, AWM, SLM, JHB

Analysis: CC, JHB, JCT,FE

Clinical expertise: AWM, SLM, EOP

Writing manuscript: CC, AWM, SLM, JHB,

Manuscript review: CC, AWM, SLM, JHB, JCT, FE

