## Supplementary Methods for "Common comorbidities in polymyalgia rheumatica and giant cell arteritis: cross-sectional study in UK Biobank"

**Supplementary Information**

Data source and study population

The UK Biobank is a large-scale multisite observational cohort study funded by the Wellcome Trust, Medical Research Council (MRC), Department of Health and Scottish governments, and Northwest Regional Development Agency. The overall study protocol (http://www.ukbiobank.ac.uk/resources/) and protocols for individual tests (http://biobank.ctsu.ox.ac.uk/crystal/docs.cgi) are available online (approval No. 5237).

Contact details for the purposes of inviting people to participate in UK Biobank were provided by the NHS, and around one in ten of those invited agreed to join the project. The assessment visit included electronic signed consent; a self-completed touch-screen questionnaire; brief computer-assisted interview; physical and functional measures; and collection of blood, urine, and saliva. A repeat assessment of 20,000 participants was carried out between August 2012 and June 2013 at the UK Biobank Coordinating Centre, Stockport, UK. Participants who lived within an approximately 30m radius of the assessment centre were invited via email to attend and undergo a repeat assessment of all the baseline measures. Data at both baseline and repeat assessment were collected using standardized proformas with the use of self-reported questionnaires [1, 2].

Eye related measurements were performed on 133 959 eligible participants at six study assessment centres and included: visual acuity, reported as Logarithm of the Minimum Angle of Resolution (log MAR), and intraocular pressure (IOP) (Goldmann-corrected; Ocular Response Analyzer, Reichert, Depew, NY, USA). Those who did not have valid IOP measurement in at least one eye were excluded from the records, leaving a total of 112 690 [3].

Definitions of cases and controls

Data on past and present conditions, obtained through a verbal interview by a trained nurse, were used in order to classify cases and controls (question code: n_20002_0_0 - n_20002_0_28 for baseline assessment and n_20002_1_0 - n_20002_1_15 for repeat assessment). All the individuals who at either of the two assessments (baseline or repeat assessment visit) reported a diagnosis of PMR or GCA (PMR code=1377; GCA code=1376) were considered as cases. All the other participants, who did not report ever having been diagnosed with PMR or GCA, were defined as controls. In addition, data about alcohol consumption, mental health, physical measures, self-reported medical conditions, health outcomes, and primary demographics were used from the baseline assessment for all cases who reported a previous diagnosis of PMR or GCA by the time of baseline assessment. For those cases who reported having been diagnosed with PMR or GCA during the repeat assessment visit, but not at the baseline assessment, only data reported at the repeat assessment were used in this analysis. For controls, the data reported during the baseline assessment were used.

Criteria for matched case-control study

A case-control study after matching cases with controls by age, sex, ethnic background and assessment centre was therefore conducted. Ethnicity was self-reported and categorised into white British, white Irish, other white background, south Asian, black (Caribbean or African), Chinese, mixed or other, Indian, Pakistani or other south Asians (including Bangladeshi) and black into Carribean, African or other black. For the purpose of this analysis, participants were classified into four categories of ethnicity: White, Black, Asian and mixed ethnic background. Cases were matched with controls from the same ethnic category, given that the prior literature states that PMR and GCA are both commoner in individuals of Northern European ancestry [4-7]. Cases were matched with controls from the same assessment centre in order to reduce any differences which might relate to health centre guidelines for diagnosis and particular centre interview practice [8]. Each case was matched by age, sex, assessment centre and ethnicity with ten controls who were not diagnosed with PMR and/or GCA in either of the two assessment periods (baseline or follow up assessment). The large sample size of UK Biobank enabled the matching of ten controls for each case with the same characteristics for all matching variables, apart from 11 cases of PMR from non-white ethnic background who were matched with controls of a different age. Eight of these PMR cases were matched with controls with age +/- 2 years, two of them with +/- 5 years and one of them +/- 10 years. Figure 1 show further details about the sample size of the matched case-control subsets. This number of controls was chosen as only a slight increase in power is obtained with the inclusion of more than 10 controls per case (Supplementary materials: Table 1-2). In addition, it would be difficult to identify more controls with the same matching characteristics from non-white ethnic backgrounds.

i)
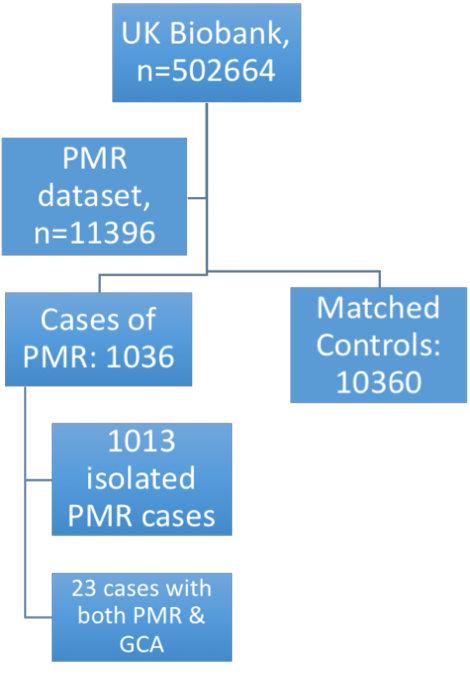
ii)
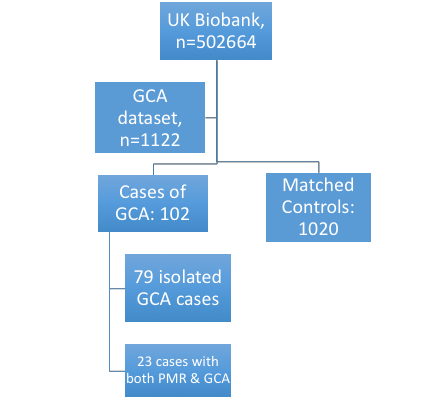

Supplementary Figure 1: Flow diagrams of the sample size for the matched case-control studies with PMR case-control dataset (i) and GCA case-control dataset (ii), matching criteria for both studies were age, sex, ethnicity and assessment centre. Matched control group includes individuals unaffected by PMR or GCA. In this figure, “isolated PMR/GCA” refers only to presence or absence of the other disease, and does not exclude participants also reporting RA or OA.

Supplementary Table 1: Power calculation for a matched PMR case-control study (1:10) based on 1036 PMR cases, assumed a 5% significance level

| OR  p0 | 1.2 | 1.4 | 1.6 |
| --- | --- | --- | --- |
| 0.2 | 0.64 | 0.99 | 100 |
| 0.4 | 0.79 | 100 | 100 |
| 0.6 | 0.77 | 100 | 100 |
| 0.8 | 0.57 | 0.97 | 100 |
| p0 probability of exposure among controls; OR Odds Ratio; power calculation for a matched case-control study calculated using STATA 14.0. | | | |

| OR  p0 | 1.2 | 1.4 | 1.6 | 1.8 |
| --- | --- | --- | --- | --- |
| 0.2 | 0.12 | 0.31 | 0.53 | 0.72 |
| 0.4 | 0.14 | 0.37 | 0.62 | 0.81 |
| 0.6 | 0.13 | 0.33 | 0.55 | 0.74 |
| 0.8 | 0.09 | 0.20 | 0.33 | 0.47 |
| p0 probability of exposure among controls; OR Odds Ratio; power calculation for a matched case-control study calculated using STATA 14.0. | | | | |

Supplementary Table 2: Power calculation for a matched GCA case-control study (1:10) based on 102 GCA cases, assumed a 5% significance level

Supplementary Table 3. Frequency of the 21 concurrent diseases that were explored in the unmatched analysis of PMR and/or GCA cases, that were selected as potential PMR/GCA mimicking conditions.

| **Comorbidities** | Unmatched Controls  (n=10031) | Cases of PMR (n=1036) | Cases of GCA (n=102) |
| --- | --- | --- | --- |
| Osteoarthritis, n (%) | 875 (8.7%) | 168 (16.2%) | 19 (18.6%) |
| Rheumatoid arthritis, n (%) | 114 (1.1%) | 38 (3.7%) | <5 |
| Spine arthritis/ Cervical spondylosis, n (%) | 145 (1.4%) | 27 (2.6%) | 5 (4.9%) |
| Psoriasis, n (%) | 131(1.3%) | 8 (0.8%) | <5 |
| Fibromyalgia, n (%) | 22 (0.2%) | 6 (0.6%) | <5 |
| Sjogren’s syndrome, n (%) | 10 (0.1%) | 5 (0.5%) | <5 |
| Peripheral neuropathy, n (%) | 23 (0.2%) | 5 (0.5%) | <5 |
| Carpal tunnel syndrome, n (%) | 13 (0.1%) | 5 (0.5%) | <5 |
| Systemic lupus erythematosus, n (%) | 11(0.1%) | <5 | <5 |
| Parkinson’s disease, n (%) | 20(0.2%) | <5 | <5 |
| Sarcoidosis, n (%) | 23 (0.2%) | <5 | <5 |
| Trapped nerve, n (%) | 32 (0.3%) | <5 | <5 |
| Psoriatic arthropathy, n (%) | 21 (0.2%) | <5 | <5 |
| Tendonitis, n (%) | 7 (0.1%) | <5 | <5 |
| Connective tissue disorder, n (%) | <5 | <5 | <5 |
| Polymyositis, n (%) | <5 | <5 | <5 |
| Peripheral nerve disorder, n (%) | 7 (0.1%) | <5 | <5 |
| Osteomyelitis, n (%) | 23 (0.2%) | <5 | <5 |
| Neck problem/injury, n (%) | 7 (0.1%) | <5 | <5 |
| Scleroderma, n (%) | 7 (0.1%) | <5 | <5 |

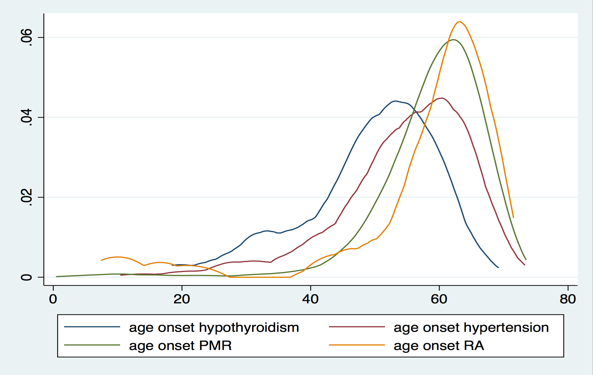

Supplementary Figure 2: Graphical representation of the age of self-reported onset of hypothyroidism, hypertension, PMR and RA among PMR cases, n=1036

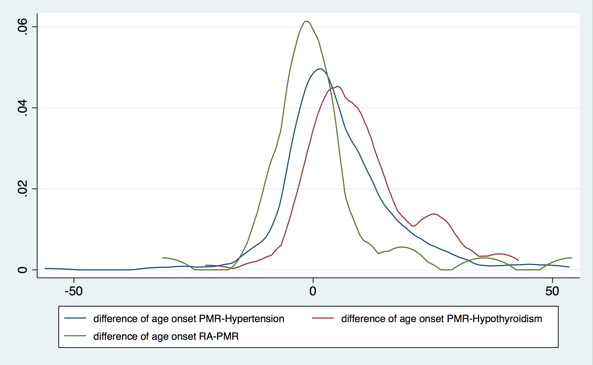

Supplementary Figure 3: Graphical representation of the mean difference between i) the age of PMR onset and age of hypertension onset, ii) the age of PMR onset and age of hypothyroidism onset, iii) the age of RA onset and age of PMR onset. On the x-axis, values less than zero indicate that the comorbidity was diagnosed after PMR; values greater than zero indicate that the comorbidity was diagnosed before PMR.

Supplementary Table 4: Frequency of recent low mood reported by PMR cases compared to matched controls

|  | PMR dataset | | | | Isolated PMR cases  vs CTRL | | | | Isolated PMR cases, receiving glucocorticoid treatment | | | | Isolated PMR cases , not receiving glucocorticoid treatment | | | |
| --- | --- | --- | --- | --- | --- | --- | --- | --- | --- | --- | --- | --- | --- | --- | --- | --- |
|  | PMR (n=1036) | CTRL (n=10360) | OR (CI) | P | PMR  (n=825) | CTRL  (n=8250) | OR (CI) | P | PMR  (n=493 | CTRL  (n=4930) | OR (CI) | P | PMR  (n=332) | CTRL  (n=3320) | OR (CI) | P |
| **Frequency of depressed mood in the last 2 weeks, n (%)*** | | | | | | | | | | | | | | | | |
| Not at all | 738 (75.0%) | 7995 (80.8%) | 1.00 | Reference | 593  (75.3%) | 6352  (80.6%) | 1.0 | Reference | 358  (75.8%) | 3802  (80.6%) | 1.0 | Reference | 235  (74.6) | 2550  (80.6%) | 1.0 | Reference |
| Several days | 193 (19.6%) | 1534 (15.5%) | 1.35  (1.14-1.60) | 5.0 x 10^-4^ | 152  (19.3%) | 1229  (15.6%) | 1.32  (1.09-1.60) | 0.004 | 93  (19.7%) | 723  (15.3%) | 1.36  (1.06-1.73) | 0.014 | 59  (18.7%) | 506  (16.0%) | 1.26  (093-1.71) | 0.1 |
| More than half the days/ Nearly every day | 53  (5.4%) | 371  (3.7%) | 1.55  (1.14-2.09) | 0.004 | 42  (5.3%) | 302  (3.8%) | 1.50  (1.07-2.10) | 0.019 | 21  (4.4%) | 194  (4.1%) | 1.16  (0.72-1.84) | 0.5 | 21  (6.7%) | 108  (3.4%) | 2.14  (1.30-3.52) | 0.003 |
| The missing category was excluded in calculation of the percentages  SD: Standard deviation; n: sample size; CTRL: number of matched controls; CI: 95% Confidence interval; OR: odds ratio; *Variables with this sign contain missing data <5%; For this analysis cases were matched with controls with the same age, sex, ethnic background and place of residence | | | | | | | | | | | | | | | | |

Supplementary Table 5: Self-rating of general health of patients with GCA compared to matched controls

|  | **GCA dataset** | | | |
| --- | --- | --- | --- | --- |
|  | GCA (n=102) | CTRL (n=1020) | OR (CI) | P |
| **Self-rating of general health, n (%)*** | | | | |
| Excellent | <5 | 152 (14.9%) | 0.42 (0.12-1.39) | 0.1 |
| Good | 28 (27.4%) | 595 (58.3%) | 1.00 | Reference |
| Fair | 41 (40.2%) | 218 (21.4%) | 4.07 (2.45-6.77) | 6.4 x 10^-8^ |
| Poor | 30 (29.4%) | 48 (4.7%) | 11.84 (6.51-5.64) | 2.2 x 10^-16^ |
| **Frequency of low mood in the last 2 weeks, n (%)*Ψ** | | | | |
| Not at all | 59 (60.8%) | 771 (80.6%) | 1.00 | Reference |
| Several days | 26 (26.8%) | 145 (15.1%) | 2.29 (1.39-3.78) | 0.001 |
| More than half the days/ Nearly every day | 12 (12.4%) | 41 (4.3%) | 3.80 (1.89-7.65) | 1.8 x 10^-4^ |
| **Number of non-cancer illnesses, n (%)*Ψ** | | | | |
| 0 | <5 | 223 (26.6%) | 0.11 (0.04-0.32) | 4.8 x10^-05^ |
| 1 | 10 (9.8%) | 213 (25.4%) | 0.29 (0.14-0.60) | 8.0 x 10^-04^ |
| 2-3 | 41 (40.2%) | 262 (31.3%) | 1.00 | Reference |
| >=4 | 47 (46.1%) | 139 (16.6%) | 2.47 (1.49-4.09) | 4.5 x 10^-04^ |
| **Ψ** The missing category was excluded in calculation of the percentages  SD: Standard deviation; n: sample size; CTRL: number of matched controls; CI: 95% Confidence interval; OR: odds ratio; *Variables with this sign contain missing data <5%; For this analysis cases were matched with controls with the same age, sex, ethnic background and place of residence | | | | |

Supplementary Table 6: Eye related measurements in PMR and cases with and without glucocorticoids at the time of the assessment

|  | PMR cases  + GC  Mean (SD) | PMR cases  – GC  Mean (SD) | GCA cases  + GC  Mean (SD) | GCA cases  - GC  Mean (SD) |
| --- | --- | --- | --- | --- |
| **Intraocular pressure** | | | | |
| Right eye | 17.3 (4.8), n=157 | 16.15 (3.6), n=108 | 18.35 (6.0), n=19 | 15.34 (3.0), n=16 |
| Left eye | 17.2 (4.7), n=156 | 15.62 (3.5), n=108 | 18.28 (6.6), n=19 | 14.87 (3.9), n=16 |
| **Visual acuity** | | | | |
| logMAR in right eye | 0.05 (0.2),  n=166 | 0.01(0.2),  n=116 | 0.07 (0.3), n=19 | 0.08 (0.2),  n=17 |
| logMAR in left eye | 0.06 (0.2),  n=165 | 0.03 (0.2),  n=115 | 0.12 (0.2), n=20 | 0.07 (0.3),  n=17 |
| **Proportion with low visual acuity (logMAR** > 0.3) in either eye | | | | |
| Low vision acuity | 25 (15.0%),  n=167 | 14 (12.7%), n=110 | 4 (20%),  n=20 | 3 (17.6%),  n=17 |
| Normal vision acuity | 142 (85.0%),  n=167 | 102 (87.9%),  n=110 | 16 (80%),  n=20 | 13 (82.3%), n=17 |
| + GC: reporting current oral glucocorticoid treatment, - GC: not reporting current oral glucocorticoid treatment at the time of assessment; SD: standard deviation; logMAR= Logarithm of the Minimum Angle of Resolution, chart which assess visual acuity. For this analysis cases were matched with controls with the same age, sex, ethnicity category and assessment centre.  *Eye related measurement were available only for 133 959 eligible participants from six study assessment centres. This is the reason why presented data contains only a small number of cases in comparison with the number of PMR and GCA cases that participate in UK Biobank* | | | | |
